# Trikafta therapy alters the CF lung mucus metabolome reshaping microbiome niche space

**DOI:** 10.1101/2021.06.02.21257731

**Authors:** Lauren M. Sosinski, Christian Martin H, Kerri A. Neugebauer, Lydia-Ann J. Ghuneim, Douglas V. Guzior, Alicia Castillo-Bahena, Jenna Mielke, Ryan Thomas, Marc McClelland, Doug Conrad, Robert A. Quinn

**Affiliations:** Department of Biochemistry and Molecular Biology, Michigan State University, East Lansing, MI, USA; Department of Microbiology and Molecular Genetics, Michigan State University, East Lansing, MI, USA; Spectrum Health, Grand Rapids, MI, USA; Department of Medicine, University of California San Diego, La Jolla, CA; Department of Pediatrics and Human Development, Michigan State University, East Lansing, MI, USA

**Keywords:** Cystic Fibrosis, sputum, microbiome, metabolome, Trikafta

## Abstract

**Background:** Novel small molecule therapies for cystic fibrosis (CF) are showing promising efficacy and becoming more widely available since recent FDA approval. The newest of these is a triple therapy of Elexacaftor-Tezacaftor-Ivacaftor (ETI, Trikafta®). Little is known about how these drugs will affect polymicrobial lung infections, which are the leading cause of morbidity and mortality among people with CF (pwCF).

**Methods:** we analyzed the sputum microbiome and metabolome from pwCF (n=24) before and after ETI therapy using 16S rRNA gene amplicon sequencing and untargeted metabolomics.

**Results:** The lung microbiome diversity, particularly its evenness, was increased (p = 0.044) and the microbiome profiles were different between individuals before and after therapy (PERMANOVA F=1.92, p=0.044). Despite these changes, the microbiomes were more similar within an individual than across the sampled population. There were no specific microbial taxa that were different in abundance before and after therapy, but collectively, the log-ratio of anaerobes to classic CF pathogens significantly decreased. The sputum metabolome also showed changes due to ETI. Beta-diversity increased after therapy (PERMANOVA F=4.22, p=0.022) and was characterized by greater variation across subjects while on treatment. This significant difference in the metabolome was driven by a decrease in peptides, amino acids, and metabolites from the kynurenine pathway. Metabolism of the three small molecules that make up ETI was extensive, including previously uncharacterized structural modifications.

**Conclusions:** This study shows that ETI therapy affects both the microbiome and metabolome of airway mucus. This effect was stronger on sputum biochemistry, which may reflect changing niche spaces for microbial residency in lung mucus as the drug’s effects take hold, which then leads to changing microbiology.

**Funding:** This project was funded by a National Institute of Allergy and Infectious Disease Grant R01AI145925

## Introduction

Cystic fibrosis (CF) is an autosomal recessive disorder caused by mutations in the cystic fibrosis transmembrane conductance regulator (CFTR) gene. CFTR is a cAMP-regulated ion channel used for the transport anions across epithelial cells *(1)*. Mutations in this gene cause a thickening of mucosal secretions, primarily in the respiratory and gastrointestinal systems, and chronic polymicrobial infection of the airways *(1)*. Common clinical representations of this disease include, but are not limited to, increased polymicrobial infections, infertility, decreased lung function, and pancreatic insufficiency *(1)*. Pancreatic sufficiency is inherently linked to the specific mutation class, but other aspects of CF pathology have unclear links to genotype *(2, 3)*. It is clear however, that those with severe disease, especially the delF508 mutation, are plagued by chronic lung infection throughout their lifetime *(4)*.

The lung microbiome of people with CF (pwCF) has been well characterized and includes bacteria, viruses, and fungi *(5–7)*. Studies of sputum expectorated from the airways have demonstrated that the CF lung microbiome diversity decreases as the disease progresses over time, becoming dominated by opportunistic pathogens, such as *Pseudomonas aeruginosa (4, 8)*. Thickened mucus within the lungs allows for these pathogens to form a biofilm and thrive *(9)*. The chemical composition of this matrix has been shown to mainly include DNA, amino acids, peptides, antibiotics, inflammatory lipids, and a myriad of small molecules from host, microbial, and xenobiotic sources *(10–14)*.

In November 2019, a new triple therapy drug Elexacaftor-Tezacaftor-Ivacaftor (ETI, Trikafta®) was approved by the United States Food and Drug Administration (FDA) for the treatment of CF *(15)*. ETI is composed of three different compounds: Tezacaftor, Elexacaftor, and Ivacaftor *(16)*. People with at least one copy of the F508del mutation, which is the most common mutation across CF patients, are eligible to take ETI. Studies from clinical trials and data available since approval have shown that the treatment is providing remarkable improvements in lung function and other disease symptoms *(17, 18)*. Little is known, however, about how this new therapy will affect the CF lung microbiome and metabolome.

In this study, sputum samples from pwCF (n=24) were collected before and after therapy (within one year of FDA approval) and analyzed using an integrated multi-omics approach including 16S rRNA amplicon sequencing and LC-MS/MS untargeted metabolomics. Changes in both microbiome and metabolome were identified and indicate a significant shift in the niche space of airway mucus and its microbial occupancy from ETI therapy.

## Methods

Further detail available in supplemental methods.

### Sample Collection

Sputum samples were collected during routine clinical visits from adult pwCF (>18 years) at two separate CF clinics (patient details table S1). Samples were obtained from the most recent clinical visit prior to ETI administration and the most recent visit after ETI administration if sputum production was possible. Ethical approval for the collections at the University of California San Diego adult CF clinic was obtained from the UCSD Human Research Protections Program Institutional Review Board under protocol #160078. Institutional review board approval was also provided for the collections at the Spectrum Health adult CF clinic in Grand Rapids, MI by the Spectrum Health Human Research Protection Program Office of the Institutional Review Board under IRB #2018-438.

### DNA Extraction, qPCR, and 16S rRNA single amplicon sequencing

A Qiagen® PowerSoil® DNA extraction kit was used to extract DNA from the sputum samples following standard protocol. PCR amplification was then performed using 27F and 1492R primers targeting the bacterial 16S rRNA gene to test for DNA amplification quality. Bacterial 16S rRNA V4 amplicon sequencing was performed with primers 515f/806r on an Illumina® MiSeq® at the Michigan State University Sequencing Core. The raw sequences were processed using QIITA (qiita.ucsd.edu(*(19)*, which is driven by QIIME2 algorithms *(20)*, and quality filtered to generate amplicon sequence variants (ASVs) through the deblur method *(21)*. ASVs in the microbiome data were classified as ‘classic CF pathogens’ or ‘anaerobes’ based on the methods of Raghuvanshi *et al*. (2020) and Carmody *et al*. (2018) *(22, 23)*. The specific ASVs and their classifications are available in table S2. qPCR was executed using universal 16S primers *(43)* and Applied Biosystems SYBR Green PCR Master Mix with three technical replicates were obtained for each sample. The microbiome data is publicly available at the Qiita repository under study #13507.

### Metabolomics

Organic metabolite extraction was performed by adding twice the sample volume of chilled 100% methanol, vortexing briefly, and incubating at room temperature for 2 hours. Samples were then centrifuged at 10,000 x g and the supernatant was collected. Methanolic extracts were analyzed on a Thermo Q-Exactive Hybrid Quadrupole-Orbitrap mass spectrometer coupled to a Vanquish ultra-high-performance liquid chromatography system. All raw files were converted to.mzXML format and then processed with MZmine 2.53 software *(24)*, GNPS molecular networking *(25)* and SIRIUS *(26)*. MZmine 2 parameters are available in the supplementary information (Table S3). The network job is available at https://gnps.ucsd.edu/ProteoSAFe/status.jsp?task=c700397169ff447490f764c34abb5abd and the mass spectrometry data were deposited on public repository <u>massive.ucsd.edu</u> under MassIVE ID <u>MSV000087364</u>.

### Statistical Analysis

Statistical approaches for both the microbiome and metabolome data were similar, due to the inherent structural similarity of the multivariate data sets. Normality of the different quantitative measures was first tested using a Shapiro-Wilk (SW) test in order to determine appropriate statistical methods. If the data was normally distributed, a paired dependent means t-test (DM t-test) was used. Otherwise, a Wilcoxon signed-rank test (WSRT) was used. Alpha-diversity was calculated for both datasets using the Shannon index. Beta-diversity measures were calculated using the weighted UniFrac distance for the microbiome and Bray-Curtis distance for the metabolome. Beta-diversity was visualized for both datasets using principal coordinates analysis (PCoA) and the EMPeror software *(27)*. Beta-diversity clustering significance pre- and post-ETI were tested using a Permutational Multivariate Analysis of Variance (PERMANOVA) method with 999 permutations. Cross population beta-diversity comparisons were done between pwCF before ETI therapy, after therapy, across the whole dataset, and within individuals pre- and post-therapy.

To identify metabolite and microbial drivers of the difference pre- and post-therapy, a random forest (RF) machine learning approach was used via the randomForest package in R *(28)*. The top 50 variables of importance were further explored. As all individual metabolite and microbiome abundance data were not considered normally distributed, statistical significance for individual microbial and metabolite changes before and after ETI treatment were calculated using the WRST. The p-values were adjusted for multiple comparisons using the Benjamini-Hochberg method.

Microbe and metabolite association vectors were calculated using mmvec *(29)*. Detail of the mmvec paramaters and analysis is available in the online supplmement.

## Results

### Microbiome and Metabolome Diversity Changes from ETI Treatment

Shannon diversity and number of ASVs of the microbiome showed an increase between samples collected before and after ETI treatment, but this did not reach statistical significance (Shannon SW normality p=0.238, WSRT p=0.062; Number of ASVs SW normality p=0.0043, DM t-test p=0.12, Fig. 1a). The Pielou evenness, however, was significantly higher post-therapy than before ETI (SW normality p=0.074, WSRT p=0.044). The metabolome did not show a significant change in Shannon index after ETI therapy (SW p=0.0017, DM t-test, p=0.45, Fig. 1b) or evenness (Pielou evenness SW p=0.13, DM t-test p=0.30). However, the number of molecular features did decrease after therapy (SW p=0.0027 DM t-test p=0.010). Collectively, this alpha-diversity analysis demonstrates that new ASVs were not being introduced into the sputum microbiome, but rather became more even with previously present taxa. In regard to the alpha-diversity of the metabolome, there was a decrease in the total number of metabolites present.

**Fig. 1.**
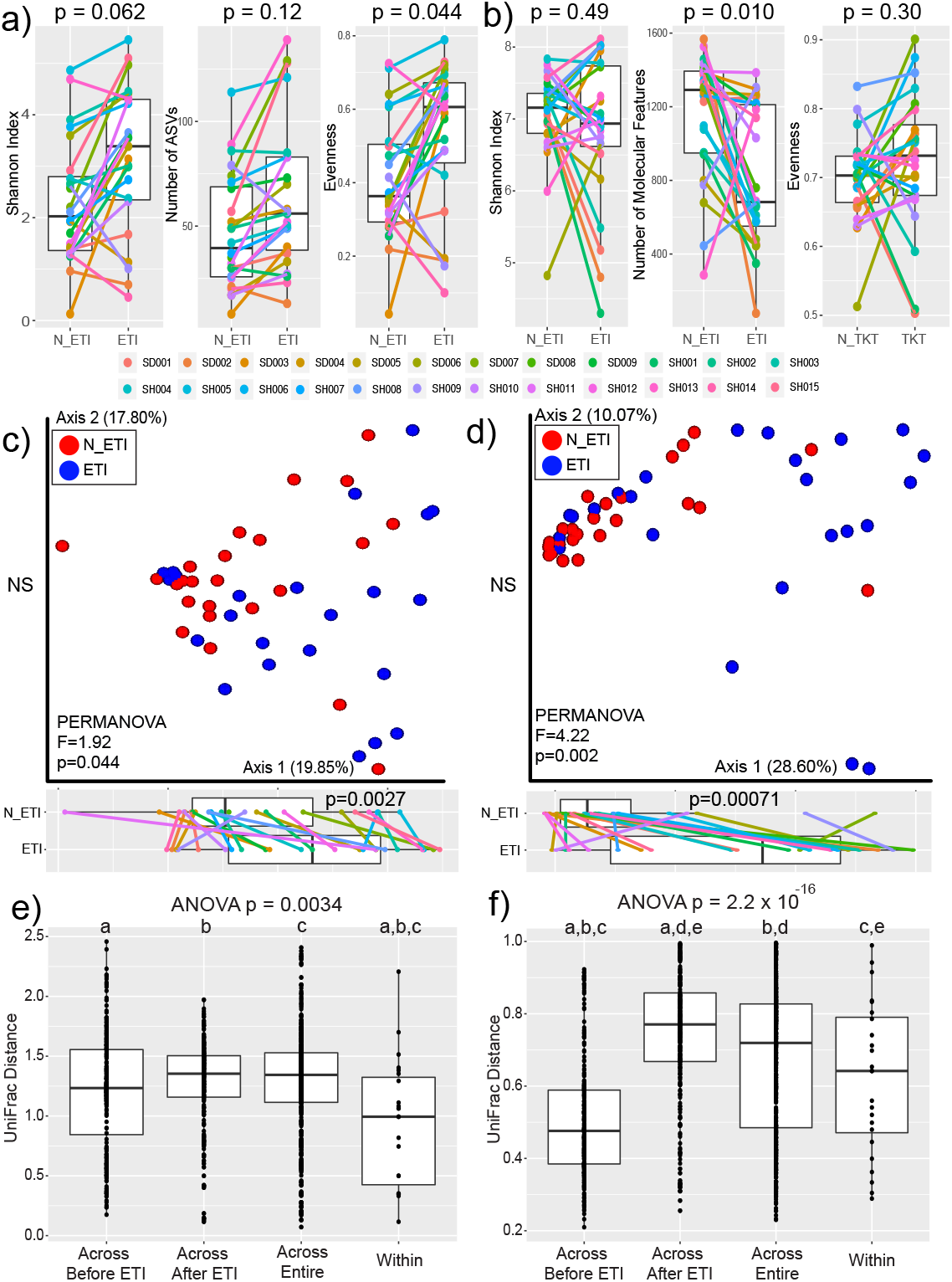
Alpha and Beta-diversity of Lung microbiomes before and after ETI. Alpha-diversity measures of a) microbiome data and b) metabolome data before (N_ETI) and after ETI therapy. P-values shown are from either the DM t-test or WSRT after testing for normality. Principal coordinate analysis plots of beta-diversity data for c) microbiome data with significance calculated utilizing the weighted UniFrac distance and d) metabolome data with significance calculated utilizing the Bray-Curtis distance. PERMANOVA statistics and the percent of variance explained by each axis are shown. Boxplots of positions on the first principal coordinate are shown tested for significance with the DM t-test. Beta-diversity cross comparisons within the e) microbiome and f) metabolome data. Cross comparisons were done across patients before and after ETI therapy, across the entire dataset, and within subjects before and after ETI therapy (within). Statistical significance was first tested with an ANOVA followed by an ad-hoc Tukey’s test. Shared letters denote distributions that are significantly different from each other.

PCoA plots were used to visualize beta-diversity differences between samples of the two data types (Fig. 1c, d). In both the microbiome and metabolome, there was a cluster of more similar samples and a spread along the first and second axis indicating samples with greater diversity. PERMANOVA testing showed that the microbiome profile of the sputum samples changed after ETI treatment (p=0.044). The metabolome profile also changed significantly after treatment (p=0.002), with a stronger metric of difference in the metabolome compared to the microbiome (F-value=1.92 microbiome, F-value=3.12 metabolome, Fig. 1c, d). There was statistically significant movement along the first principal coordinate axis after ETI therapy for both the microbiome (SW test p=0.02, DM t-test p=0.0027) and the metabolome (SW p=0.0011, DM t-test p=0.00071) indicating that changes in these measures occurred similarly within this beta-diversity space. There was no significant change along the second axis. The beta-diversity differences among samples were also compared before, between, and after ETI treatment across subjects and within subjects (SW normality on beta-diversity distributions microbiome p=1.4 × 10^−15^, metabolome p=2.2 × 10^−16^). The microbiome showed the smallest change within subjects before and after therapy than across subjects at any period, indicating that although the microbiome profiles change significantly (Fig. 1e), there was still more similarity within an individual before and after therapy than across the population no matter the treatment category. The metabolome beta-diversity comparisons showed different trends than the microbiome. The largest beta-diversity in metabolite profiles was seen across patients in samples collected after therapy, signifying that the chemical makeup of sputum becomes far more varied across people once administered ETI (Fig. 1f). In contrast, the metabolomes were the most similar across subjects prior to ETI therapy, indicating that sputum metabolite profiles were relatively similar prior to ETI administration, but varied greatly across individuals after treatment.

### Microbial Changes After ETI Treatment

A random forest classification was used to determine how well the microbiome data reflected the pre- or post-treatment groups and to rank the ASVs by their contribution to that classification. Overall, the random forest model poorly classified the microbiome data with an error rate of 44.7%. *Veillonella parvula* and *Staphylococcus* sp. were ASVs with strong classifiers (Table S4) with a high overall abundance. However, none of the ranked ASVs were significantly different between pre- and post-treatment samples after correction for false-discovery (Benjamini-Hochberg corrected, WSRT p>0.05). The ASV representing *Pseudomonas* showed dynamic changes in some individuals, but it was not significantly different in the overall paired data (Fig. 2). Similarly, at the family level, there was no significant difference before and after therapy following false discovery rate correction. We also summed the abundance of all ‘CF pathogens’ and ‘anaerobes’ (as described in table S2) and compared the log-ratio of pathogens/anaerobes, as described by Raghuvanshi *et al*. (2020) *(22)*, and found it significantly decreased following ETI therapy (SW p = 0.233, WSRT p = 0.013, Fig. 2).

**Fig. 2.**
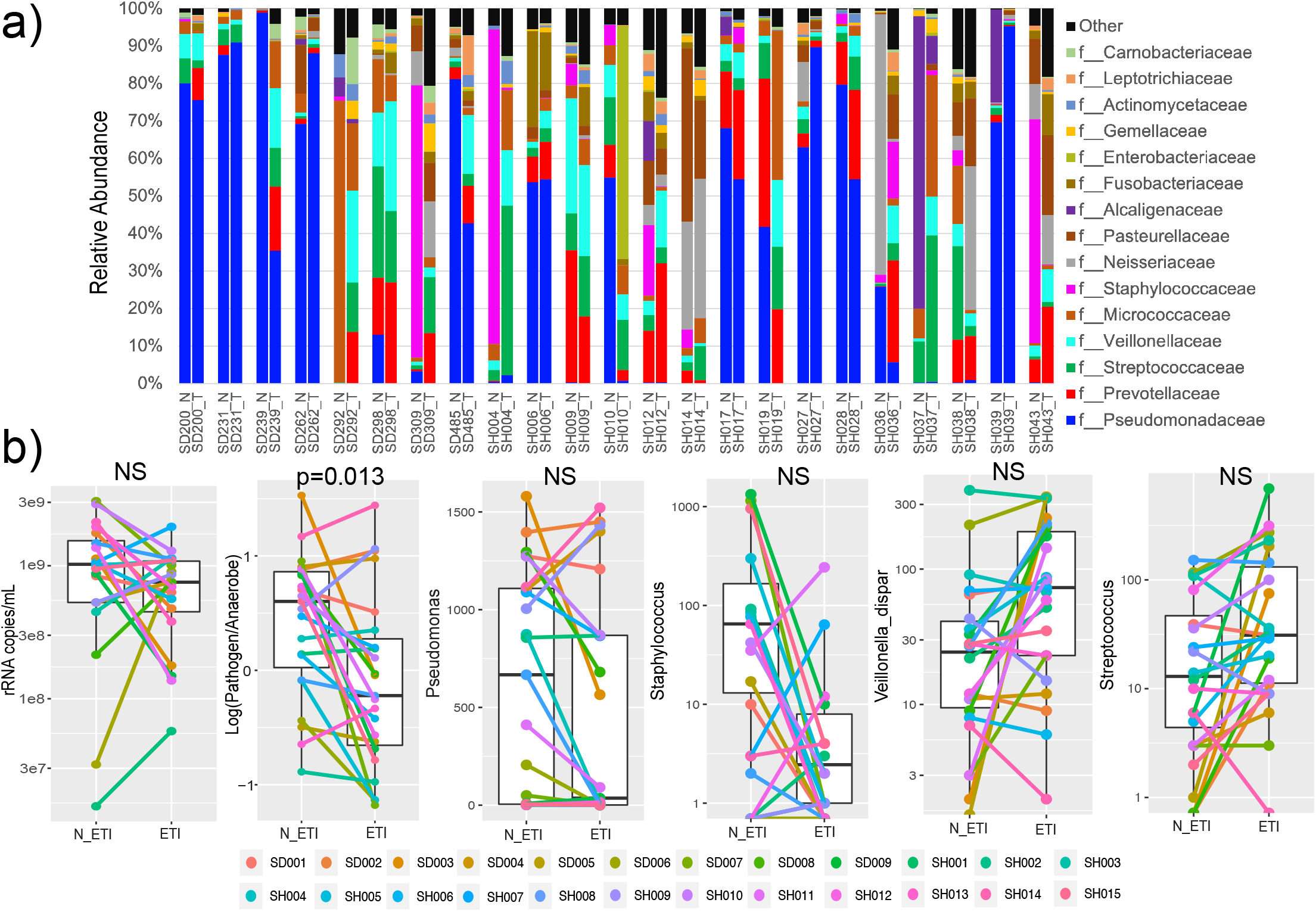
Microbiome changes throughout ETI therapy. a) Taxonomic dynamics at the family level of ASVs in each subject before (N) and after (T) ETI therapy. b) The rRNA copies/mL of sputum, log-ration of pathogen:anaerobes and ASV dynamics before and after ETI therapy.

A qPCR assay using universal primers for the bacterial 16S rRNA gene *(43)* was used to calculate the total number of rRNA copies/mL of sputum pre- and post-therapy. The mean prior to therapy was 1.17 × 10^9^ copies/mL and after therapy was 7.62 × 10^8^ copies/mL. This difference was not statistically significant but did show a decreasing trend (SW p = 0.0027, DM t-test p = 0.061, Fig. 2).

### Metabolite Changes After ETI Treatment

Annotated metabolites were first grouped into molecular families and summed to determine overall changes. This analysis showed the major metabolomic signature due to ETI therapy was a decrease in peptides and amino acids. Though phosphocholine and phosphoethanolamine abundance did not change before and after ETI, peptide and amino acid abundance significantly decreased (Fig. 3a). This demonstrates an overall change in the relative abundance of this compound class, while other classes remained static.

**Fig. 3.**
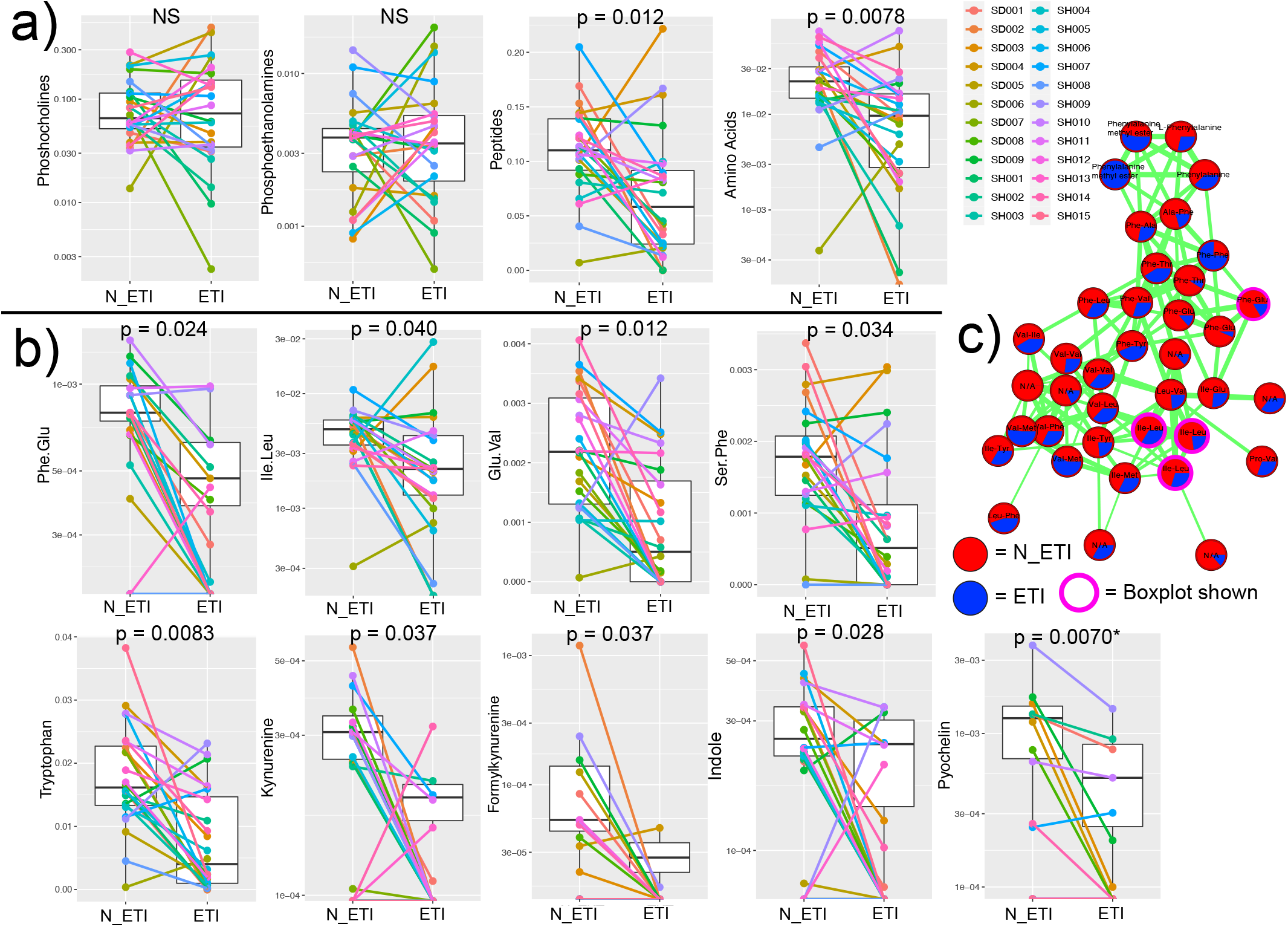
Metabolite network of peptides and other metabolite changes. a) Molecular family metabolite abundance changes pre- and post-therapy. b) Individual metabolite changes pre- and post-therapy. c) Molecular network of peptides identified by GNPS library searching. Each node represents a unique MS/MS spectrum (putative metabolite), connections between the nodes are determined and width-scaled by the cosine score from MS/MS alignment. Pie charts are the total feature abundance colored according to the legend.

A random forest machine learning classification was used to assess how well the complete metabolomic data reflected changes after ETI therapy. The out-of-bag error rate of the classification was 22.92% indicating that there was a metabolomic signal for ETI therapy, but not all samples were correctly classified as pre- or post-treatment. The importance of each metabolite in the construction of the random forest model was produced, noting the impact of each metabolite on differentiating pre- and post-treatment groups (Table S5). Of the 50 most important classifiers, 13 were annotated in the GNPS database and of those, 10 were annotated as amino acids or peptides. These were primarily dipeptides, including Phe-Glu, Ile-Leu, Glu-Val, and Ser-Phe, as well as the amino acid tryptophan; all of which significantly decreased after ETI therapy (Fig 1b). Molecular network analysis (Fig. 3c) showed a diverse set of peptides that were more abundant prior to ETI therapy. Metabolites from the kynurenine pathway (which includes tryptophan) were also identified as strong classifyers in the model. Kynurenine, formylkynurenine, and indole abundance significantly decreased after ETI therapy (Fig. 3b). The *P. aeruginosa* siderophore pyochelin was detected in only 10 of the 24 patients. Comparing pyochelin abundance within those 10 individuals showed that it significantly decreased after ETI therapy as well (Fig. 3b). Though commonly detected in CF sputum with the metabolomics methods used here, other *P. aeruginosa* specialized metabolites were not detected in this study, except for one quinolone (NHQ) that was detected in 6 samples.

### Trikafta Metabolism in CF Mucus

ETI is a triple therapy of the compounds Ivacaftor, Elexacaftor, and Tezacaftor. All three drugs were identified in the sputum metabolome by MS/MS analysis with similar fragmentation behavior to that described by Reyes-Ortega *et al.* (2020) *(30).* This included the known and unknown metabolized products of the parent drugs with related MS/MS spectra (Fig. 4). Ivacaftor had extensive metabolism revealed by molecular networking with the parent drug having six related nodes with unique retention times. Two of these are known, M1 and M6, as hydroxymethyl ivacaftor and ivacaftor carboxylate, respectively. Other modifications of the compounds were also seen, including further hydroxylations and carboxylations. Tezacaftor metabolism was also identified. This included dehydrogenation (metabolite M1, *m/z* 519.1400, C_26_H_26_F_3_N_2_O_6_+H^+^). Also detected, but not shown, was a phosphorylated metabolite (*m/z* 599.1401, C_26_H_27_F3N_2_O_9_P+H^+^) and a known glucuronate (metabolite M3, Fig. 4a). Elexacaftor exhibited only one metabolic transformation – the loss of a methyl group on its pyrazole ring (Fig. 4).

**Fig. 4.**
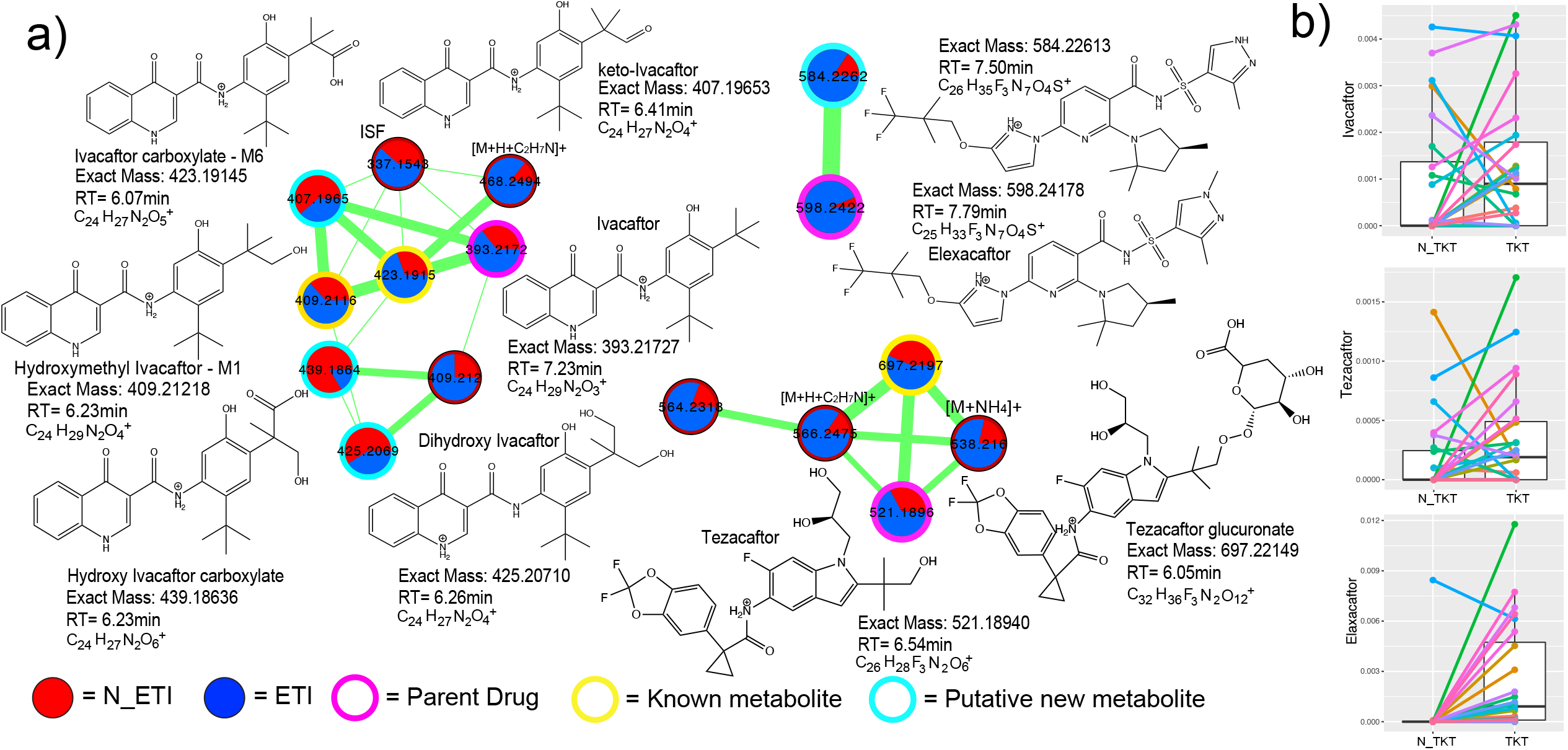
ETI metabolism detected in sputum metabolomic data. a) Three separate molecular networks are shown for Ivacaftor, Tezacaftor, or Elexacaftor and their related metabolic products as identified by MS/MS spectral alignments. Each node in a network represents a unique MS/MS spectrum and connections between the nodes indicate spectral similarity as identified by the cosine score. The width of the edges are scaled to the cosine score and the pie chart inside nodes represent the sum of the area-under-curve abundance of that molecule in either pre-(red) or post-treatment (blue) sputum samples. The nodes are highlighted by whether they represent parent drug, known metabolized product, or putative unknown metabolized product. Putative structures of the metabolites are shown with their molecular formulas, retention times, and exact masses. Note that the stereochemistry of some of the metabolized products cannot be discerned with this level of MS/MS annotation. b) Boxplots of the area-under-curve abundance of the three parent drugs in pre- and post-ETI samples.

Ivacaftor and Tezacaftor were detected in sputum samples both prior to and after ETI administration. These two compounds were released as therapies in prior formulations of CFTR correctors, likely explaining their presence. Elexacaftor, the next generation corrector unique to ETI, was present in sputum after its prescription as expected. However, one subject unexpectedly had Elexacaftor present in their lung sputum prior to clinical knowledge of administration of ETI. The presence of Ivacaftor and Tezacaftor in the sputum of pwCF prior to administration of ETI led to the investigation of whether or not the presence of previously approved correctors/potentiators in sputum prior to administration of ETI may have buffered the microbiome dynamics observed. Ivacaftor (a component of Kalydeco®, Orkambi®, and Symdeko®) was found in 11 of the 24 patients prior to ETI therapy. There was no significant difference in the alpha or beta-diversity changes between subjects that had Ivacaftor in their sputum prior to ETI and those that did not (p>0.05, Fig. S1). This indicates that prior CFTR corrector/potentiator therapy was not contributing significantly to the overall changes seen with ETI in this study, allowing these changes to be definitively attributed to this specific triple therapy

### Microbiome/Metabolite Associations Through ETI Therapy

To associate microbiome and metabolome dynamics across the dataset, we employed mmvec. This is a novel algorithm robust to the challenges of compositionality of omics datasets that calculates conditional probabilities of the association between all ASVs in the microbiome data with all metabolite features. The overall neural network showed a strong association between a changing microbiome and metabolome (Fig. S2). Additionally, the biplot of the mmvec algorithm enabled visualization of the microbiome vectors associated with the metabolomic space. In the biplot, a clear separation in vector directionality was found between pathogen and anaerobe associations with the metabolome. This indicates that the metabolites that change in association pathogens are not the same metabolites that associate with changing anaerobes. Drivers of the metabolite changes associated with pathogens were mostly peptides (Fig. 5a); those same peptides shown to be decreasing after ETI therapy. Plotting the conditional probabilities of each peptide with the mean of all pathogens and all anaerobes in the dataset showed that the peptides were significantly associated with pathogens (WSRT p<0.001) (Fig. 5b). Kynurenine, another metabolite found to decrease with ETI therapy, was most strongly associated with pathogens. This analysis indicates the decrease of peptides and kynurenine in sputum samples collected from pwCF is associated with a decrease in the relative abundance of pathogens (Fig. 5c).

**Fig. 5.**
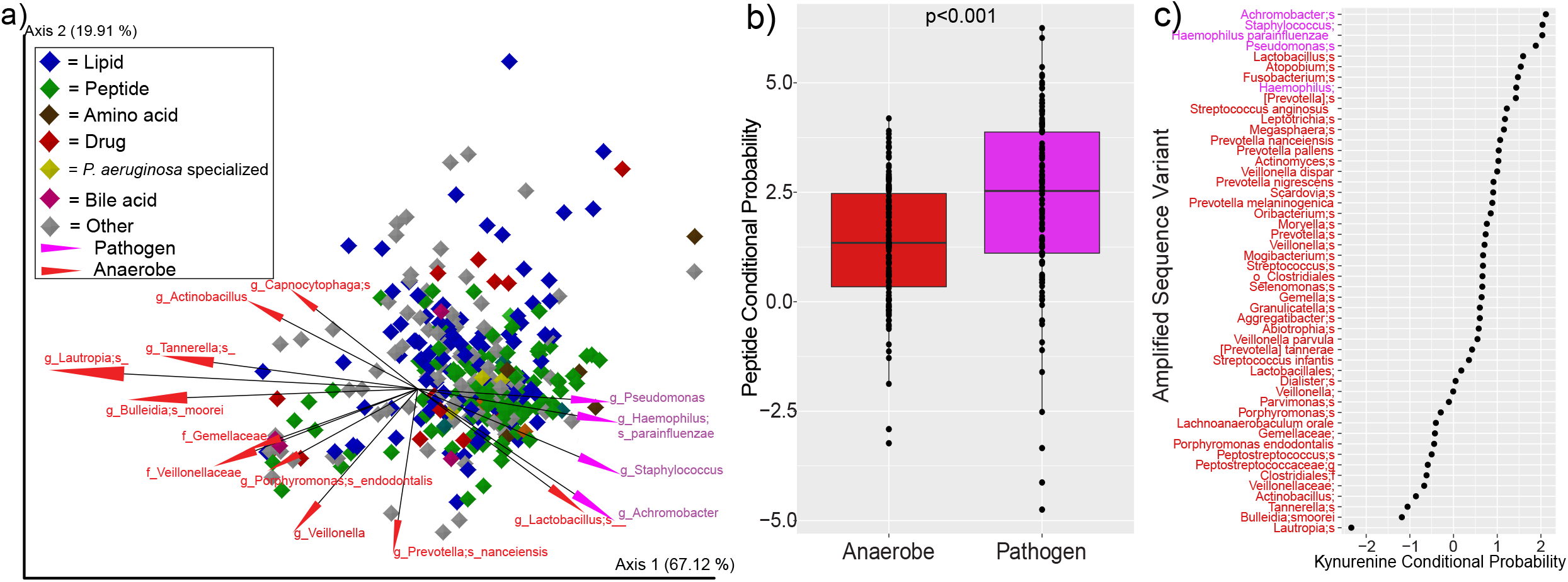
Mmvec analysis of sputum microbiomes and metabolomes from pwCF. a) Biplot of the metabolite and microbe vector associations. Each diamond represents a metabolite (only metabolites annotated within the GNPS library are shown) and they are colored by their molecular family. The vectors are the top 15 ASVs associated with the metabolomic dynamics and they are colored by whether or not they are considered clinical pathogens or anaerobes. b) Conditional probability distributions for the mean of all peptides identified in the dataset and their association with either anaerobes (red) or pathogens (purple, p-value from DM T-test). c) Rank abundance of the conditional probabilities of kynurenine with different anaerobes (red) and pathogens (purple) ASVs.

## Discussion

This study assessed the multi-omic changes in sputum from pwCF after administration of the novel CF triple therapy ETI. This promising new therapy has shown significant improvement in lung function and symptom measures of pwCF in clinical trials with great potential to improve the lives of these individuals *(15, 16)*. Improving CFTR function is a major goal of ETI therapy as it is the underlying cause of CF. However, many adult patients have been living with persistent lung infections and an evolving microbiome for decades. Promising as the treatment is, it is mostly unknown how the therapy will affect lung infections and the chemistry of sputum. This is of paramount importance; if the microbial infections in the lungs of pwCF do not clear and/or change favorably, then the full benefits of the therapy may not be realized. Preliminary studies of other CFTR modulators and correctors, specifically Ivacaftor which has received the most attention due to having the earliest FDA approval, have shown some changes in microbial diversity measures with treatment *(31)*, specifically in the gut *(32, 33)*, but most studies find little change in the airway microbiome *(33–36)*. The addition of CFTR correctors, such as Lumacaftor, have shown an increase in microbial diversity in the CF airways *(37)*, but other studies also show less marked responses *(38)*. This study, to our knowledge, is the first to look specifically at the microbiome and metabolome changes resulting from ETI therapy (which includes the new highly effective potentiator Elexacaftor). Effects of the treatment were seen in both the microbiome and the metabolome. By beta-diversity measures, the effect was stronger in the metabolome, demonstrating that the drugs were altering the biochemical environment of CF mucus.

Microbial diversity increased with ETI therapy, indicating that the microbiome in the lungs of pwCF is becoming more complex. This increase was driven by a higher microbial evenness, a metric that contributes to the Shannon diversity, though the Shannon index itself did not reach statistical significance. Therefore, the lung microbiome of pwCF was not necessarily gaining new or losing old microbial members after ETI therapy, but those present became more similar in their relative abundances, possibly an indication of greater community stability. The overall profiles of the microbiome were significantly changed after ETI therapy and did so in a similar way, as shown by the homogeneous directional movement across the first principal component axis. Despite these overall changes, there was no single organisms that significantly altered from ETI therapy after multiple-comparisons correction. This is likely due to the widely known personalization in the CF microbiome *(8, 39)*. Individuals have very different microbial profiles, so the start and endpoints from any pharmaceutical treatment may not be universal across subjects. This personalization was again observed here; subjects were still more similar to themselves after ETI therapy than to other subjects. A larger sample size in this study may have reached statistical significance for microbial taxa of interest because the trends for pathogens, such as *P. aeruginosa* and *Staphylococcus*, were showing strong reductions in abundance, while anaerobes were showing an increase. Accordingly, a collective comparison of the decrease in the log-ratio of the pathogen:anaerobe abundances did reach statistical significance. This finding validates this approach of simplifying the microbiome to these two communities and indicates that there is an overall reduction in the relative abundance of collective clinical pathogens, even though not all patients have the same pathogen. There was also a trend in the decreased bacterial load after ETI therapy, supporting that the increase in diversity seen from microbiome measures may be associated with a decrease in total bacterial load from expectorated sputum, though this too did not reach statistical significance. In summary, ETI therapy significantly altered the lung microbiome, exemplified as an increased microbial evenness driven by a reduction in the relative abundance of pathogens in place of an increase in the relative abundance of anaerobes. This increase in anaerobes may have clinical relevance for treating lung infections in the new era of highly effective CFTR modulators. The role of anaerobes within the CF lung is rather unclear as they are associated with better lung function *(40)*, but also pulmonary exacerbations *(22, 23, 41)*. In light of these findings, antibiotic treatment with more anaerobic coverage may be an effective approach to treat CF infections in those prescribed ETI.

The metabolome showed significant changes following ETI therapy, with stronger metrics than the microbiome. This was characterized by an increase in variability across subjects. Lung sputum metabolomes were relatively similar prior to ETI therapy, but, when on drug, the sputum metabolomes were highly diverse across subjects. These interesting chemical dynamics indicate that ETI induces a sort of metabolomic turmoil within the airways of pwCF, where the lung sputum biochemistry changes significantly with a highly varied outcome across individuals. However, similar to changes in the microbiome, the directionality of change was uniform, indicating a common metabolomic shift driven by ETI. Despite this variability, there were some uniform changes identified, particularly changes in peptides, amino acids, and kynurenine metabolism. The latter was identified as an important pathway associated with *P. aeruginosa* dynamics from Lumacaftor/Ivacaftor therapy in a previous study *(38)*, which the findings here and may be a universal consequence of CFTR modulator treatment. The decrease in the overall abundance of peptides, particularly dipeptides, links these metabolites to a previous study that associated them with worsening lung function *(13)*. These peptides were shown to be sourced from neutrophil elastase activity. Thus, the decrease in these metabolites with ETI therapy may also represent a reduction in this inflammatory process, though inflammatory markers were not measured in this study. There may be a link between the decrease in kynurenine metabolism and amino acids/peptides, as it is a principal pathway for the metabolism of tryptophan in humans and bacteria. The reduction of peptides in sputum may reduce their availability for pathogens, particularly *P. aeruginosa*, to metabolize through the kynurenine pathway or others. The association of kynurenine metabolism with *P. aeruginosa* in a previous CFTR modulator study supports the hypothesis that this therapeutic approach may reduce these compounds and reshape the carbon source available to the pathogen.

Microbial metabolite vector associations further supported the changing relationship between peptides, kynurenine metabolism, and pathogens. This approach, robust to the statistical challenges of cross-omics comparisons from compositional datasets *(29, 42)* in addition to identifying microbiome and metabolome changes, showed that the decrease in peptides and kynurenine was associated with a reduction in pathogens. In light of this finding, we propose the hypothesis that ETI therapy, and possibly other CFTR modulators, reshape CF microbiome niche space by reducing peptide and amino acid availability. This shift may squeeze out pathogens, which are known to preferentially metabolize amino acids in CF mucus *(43–46)*.

Metabolomics of complex clinical samples often identifies xenobiotics such as drugs administered to patients *(14)*. Prior CFTR modulators were detected in the clinical samples from pwCF in this study, including evidence of diverse metabolism of these drugs, particularly ivacaftor. This created a unique opportunity to determine whether or not the presence of a previously approved CFTR modulator therapy in a patient’s sputum affected the microbiome and metabolome dynamics of ETI. Interestingly, there was no difference in the microbial and metabolite dynamics described between those taking CFTR modulators and those not. This is evidence the significant changes described in this study were driven by ETI, perhaps Elexacftor itself, which is known to be a highly effective CFTR potentiator. These results show promise that ETI therapy may have a particularly strong effect on the CF sputum microbiome and metabolome where other CFTR modulators have not *(33–36)*.

There are several caveats to our study, perhaps most importantly, ETI therapy is known to reduce sputum production in pwCF. It is difficult to discern if the changes we identified here are due to sputum chemical and microbial dynamics or a change in the ability to produce sputum when on CFTR modulators. Instructions for expectoration were not varied before and after ETI to normalize the sampling approach and all subjects in this study were able to produce sputum for collection both prior to and during ETI therapy. Furthermore, the relatively small sample size may have masked some specific changes, particularly with individual microbial ASVs. Larger studies of pwCF before and after ETI therapy are warranted, though with the wide availability of these CFTR modulators currently makes collecting ETI naïve sample difficult. Harvesting sputum samples for CF biobanks could be an effective alternative. Finally, as with all untargeted metabolomics methods, a single protocol for metabolite extraction and mass spectrometry analysis identifies only a fraction of the total metabolite pool. Future studies on the metabolome of CF sputum in response to ETI therapy with more varied protocols may reveal other important pathways that are altered by these drugs.

This study shows that the highly effective CF triple therapy ETI induces significant changes in the CF sputum microbiome and metabolome. This is exemplified by an overall reduction in pathogens compared to anaerobes in addition to reduced amino acid availability and kynurenine metabolism with therapy. This shows promise for the future of CF infection therapeutics because if pathogens are decreasing in abundance in place of anaerobes, antimicrobial therapy can be targeted to these organisms that may have less intrinsic resistance than notorious pathogens, such as *P. aeruginosa* and *S. aureus*.

## Data Availability

The mass spectrometry data were deposited on public repository massive.ucsd.edu under MassIVE ID MSV000087364.

## Acknowledgements

The authors would like to thank the Spectrum Health and UC San Diego clinical teams who assisted with sample and metadata collection for this study. We would like to acknowledge funding from the National Institutes of Allergy and Infectious Diseases under R01 grant R01AI145925 awarded to PI Quinn.

