## Supplementary figures and images for "Trikafta therapy alters the CF lung mucus metabolome reshaping microbiome niche space"

### Figure S1

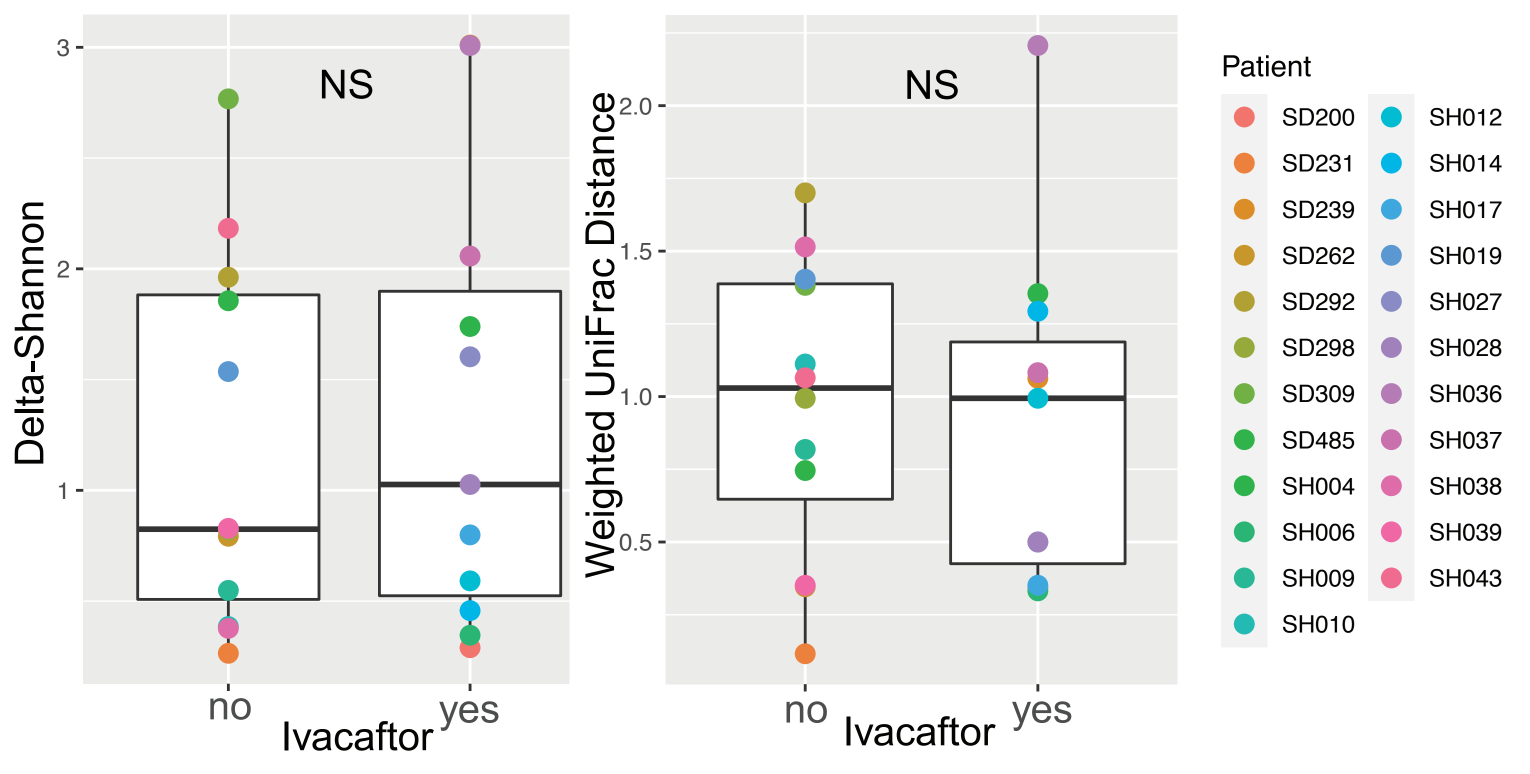

### Figure S2

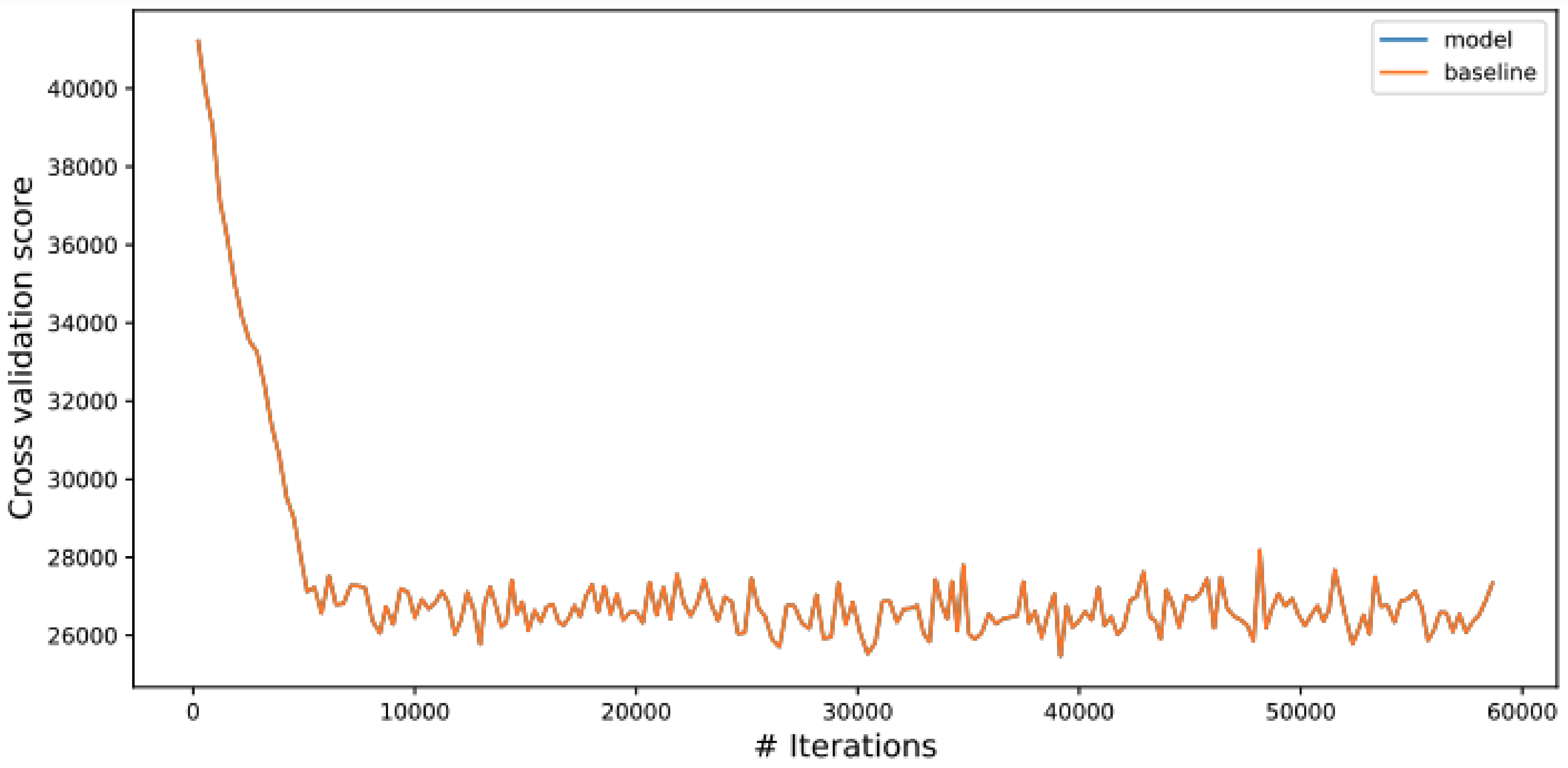

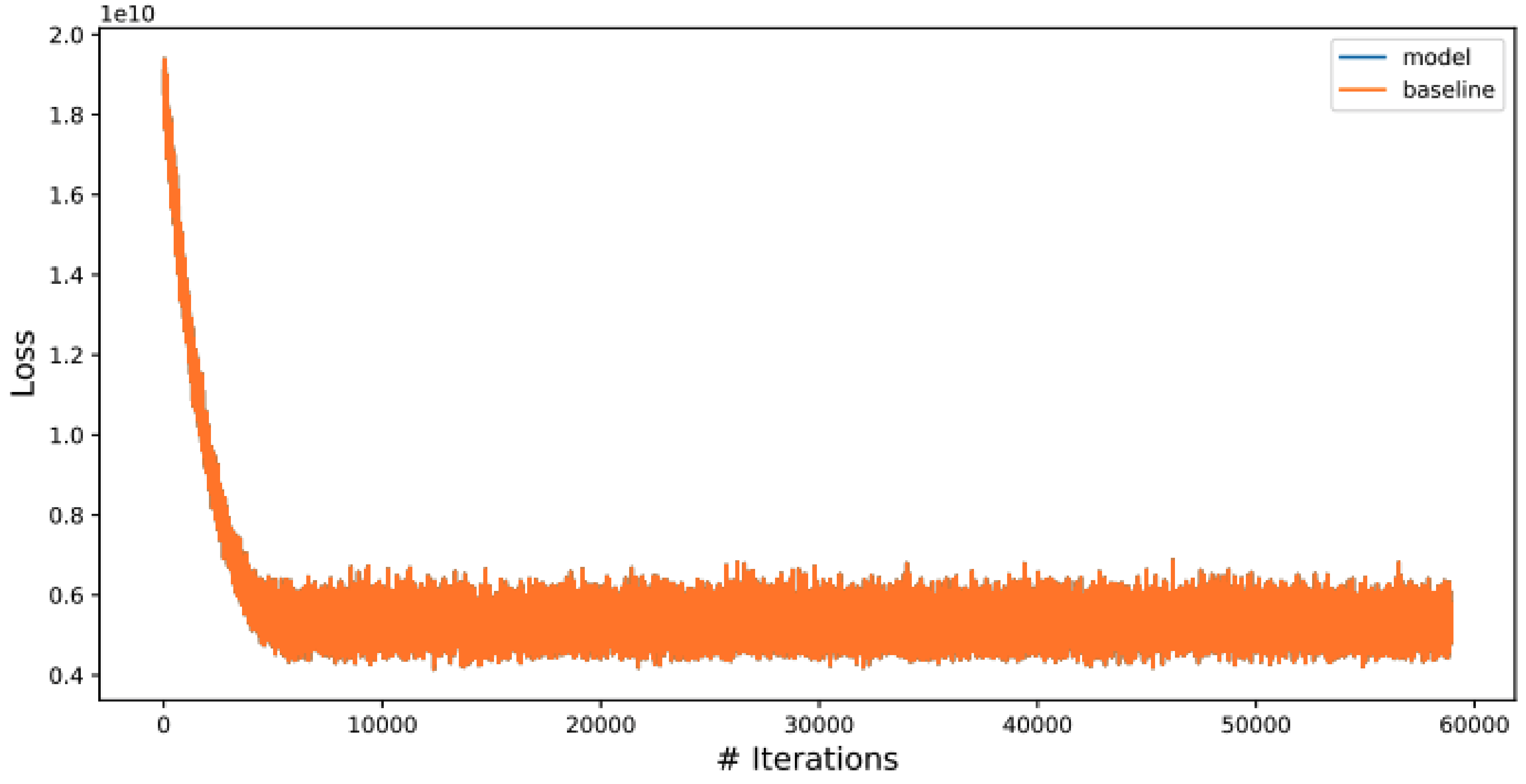
